# Spousal concordance in adverse childhood experiences and the association with depressive symptoms in middle-aged and older adults: findings across China, the US, and Europe

**DOI:** 10.1101/2022.10.28.22281641

**Authors:** Ziyang Ren, Weidi Sun, Siyu Zhu, Siqing Cheng, Wen Liu, Ho Cheung William Li, Wei Xia, Changzheng Yuan, Davies Adeloye, Igor Rudan, Dexter Canoy, Peige Song, the Global Health Epidemiology Research Group (GHERG)

**Author notes:** Corresponding Author: Peige Song, PhD, School of Public Health and Women’s Hospital, Zhejiang University School of Medicine, Hangzhou 310058, China. These authors contributed equally to this work (Ziyang Ren, Weidi Sun).

## Abstract

**Importance:** Adverse childhood experiences are associated with higher depressive risks in adulthood. Whether respondents’ adverse childhood experiences are associated with their adulthood depressive symptoms and further contribute to spousal depressive symptoms was unexplored.

**Objective:** To assess the spousal concordance of adverse childhood experiences, the association of respondents’ adverse childhood experiences with spousal depressive symptoms and the mediating role of respondents’ depressive symptoms in this association.

**Design:** This cohort study was conducted based on data from China Health and Retirement Longitudinal Study (CHARLS), the Health and Retirement Study (HRS), and the Survey of Health, Ageing and Retirement in Europe (SHARE). Data were analyzed from June through July 2022.

**Setting:** The study was based on three cohorts in China, the US, and Europe.

**Participants:** Couples aged 50 years or older with complete data on ACEs and covariates.

**Exposures:** Adverse childhood experiences.

**Main Outcomes and Measures:** The 10-item Center for Epidemiological Studies Depression Scale, the eight-item Center for Epidemiologic Studies Depression Scale, and the Europe-depression scale were respectively applied in CHARLS, HRS, and SHARE to define depressive symptoms.

**Results:** Couples’ adverse childhood experiences were noticeably correlated in the three cohorts. Significant associations between husbands’ adverse childhood experiences and wives’ depressive symptoms in the three cohorts, with ORs and 95% CIs of 2.09 (1.36-3.22) for 4 or more adverse childhood experiences in CHARLS, and 1.25 (1.06-1.48) and 1.38 (1.06-1.79) for 2 or more adverse childhood experiences in HRS and SHARE. However, wives’ adverse childhood experiences were associated with husbands’ depressive symptoms in only CHARLS and SHARE. Findings in intra-familial and extra-familial adverse childhood experiences were consistent with our main results. Additionally, respondents’ depressive symptoms mediated more than 20% of the effect of respondents’ adverse childhood experiences on spousal depressive symptoms.

**Conclusion and Relevance:** In the CHARLS, HRS, and SHARE databases, we found that adverse childhood experiences were significantly correlated between couples. Respondents’ adverse childhood experiences were associated with spousal depressive symptoms, with respondents’ depressive symptoms mediating the association. The two-way implications of adverse childhood experiences on depressive symptoms should be considered at couple level and effective interventions are warranted.

**Key Points:** *Question:* Whether respondents’ adverse childhood experiences are associated with their adulthood depressive symptoms and further contribute to spousal depressive symptoms?

*Finding:* This study found significant associations between husbands’ adverse childhood experiences and wives’ depressive symptoms. Additionally, respondents’ depressive symptoms mediated more than 20% of the effect of respondents’ adverse childhood experiences on spousal depressive symptoms.

*Meaning:* The implications of childhood adversity on later-life depressive symptoms at the couple level should be considered and strengthening the effective interventions of adverse childhood experiences and depressive symptoms is needed.

## Introduction

Depression is a common psychological disorder characterized by a persistent feeling of sadness and/or lack of pleasure^1^. According to the Global Burden of Disease Study (GBD) 2019, there were 170.8 million cases of depression in 1990 and 279.6 million cases in 2019, corresponding to an increase of 63.7%^2^. Depressive symptoms also play an important role in the development of chronic diseases and disabilities, imposing an enormous social and economic burden^3,4^.

There is a growing understanding that adverse experiences in childhood can heighten one’s risk of depression later in life. Adverse childhood experiences (ACEs) refer to a wide spectrum of intra-familial and extra-familial traumatic events occurring in childhood, such as family violence and bullying^5,6^. The deleterious impacts of ACEs on life course health and related medical burdens have received increasing attention recently^7,8^. It has been found that ACEs are significantly associated with increased risks of numerous diseases, particularly mental disorders ^9,10^.

Previous research found that depressive symptoms in older couples were correlated^11^. However, whether this correlation is caused by pre-marital or post-marital factors remains unclear. According to recent research, newly married couples were consistent in their mental health and cardiovascular risks^12,13^, both of which are potential consequences of ACEs^14,15^, suggesting that couples may have a similar history of ACEs. Furthermore, given that respondents’ ACEs have adverse implications on their spouses’ mental health^11,16^, it is reasonable to deduce that ACEs may induce respondents’ depressive symptoms and further spousal depressive symptoms. However, there is limited research exploring this hypothesis.

To fill these research gaps and verify the above hypothesis across the social development spectrum, this study used population-based datasets from three regions with different social development circumstances: the China Health and Retirement Longitudinal Study (CHARLS)^17^, the Health and Retirement Study (HRS) in the United States (the US)^18^, and the Survey of Health, Ageing and Retirement in Europe (SHARE)^19^ to investigate the spousal concordance of ACEs, the association between respondents’ ACEs and spousal depressive symptoms, and the mediating role of respondents’ depressive symptoms in this association.

## Method

### Data sources and study population

The CHARLS enrolled participants aged 45 years and above from 150 counties within 28 provinces across China, using a stratified multistage probability sampling strategy^17^. The baseline survey was conducted in 2011 and three follow-up surveys have been conducted in 2013, 2015, and 2018. Information on ACEs was additionally collected in the 2014 life history survey from all living respondents in the 2011 and 2013 surveys.

The HRS is a population-based longitudinal survey of participants aged over 50 years in the United States. More details of the study design and recruitment can be found elsewhere^18^. Respondents were followed biennially from entry in 1992 until 2018. The most updated and completed data on ACEs were collected in the 2012 survey.

The SHARE is a multidisciplinary and longitudinal study of people aged 50 years or older from 28 European countries and Israel^19^, conducted biennially from 2004 until now. Our study included participants from the latest 8^th^ wave in 2018^20,21^, and data on ACEs were from the 7^th^ wave in 2017 SHARELIFE study ^22,23^.

The CHARLS, HRS and SHARE were approved by the Institutional Review Board at Peking University, the Institutional Reviewing Board at the University of Michigan and the National Institute on Aging, and the Ethics Council of the Max Planck Society, respectively. Written informed consent was obtained from all respondents. In this study, couples aged 50 years or older with complete data on ACEs and covariates were included. The study flow charts can be found in **eFigures 1**-**3**.

### Definition of ACEs

Detailed definitions of ACEs are shown in **eTable 1**.

For the CHARLS, ACEs before the age of 17 years were assessed by dichotomous or multiple-choice questions. We identified 14 ACE events, including 11 intra-familial domains (emotional neglect, family violence, parental separation or divorce, parental substance abuse, parents incarcerated, parental mental illness, parental disability, parental death, sibling death, physical abuse, and economic adversity)^24,25^ and 3 extra-familial domains (bullying, loneliness, and community violence)^26-28^. All domains were further dichotomized and summed to obtain overall ACEs, intra-familial ACEs, and extra-familial ACEs. Overall and intra-familial ACEs were then classified as 0, 1, 2, 3, and 4 or more; extra-familial ACEs were classified as 0, 1, and 2 or more.

In the HRS, the Psychosocial and Lifestyle Questionnaire was utilized to ask about childhood traumas before the age of 18 years^29^. In this study, a total of 6 ACEs were identified, including 4 intra-familial domains (emotional neglect, parental substance abuse, physical abuse, and economic adversity) and 2 extra-familial domains (repeating school year and trouble with police)^30^. Overall ACEs, intra-familial ACEs, and extra-familial ACEs were further categorized based on the number of ACEs, with values ranging from 0 to 6, 0 to 4, and 0 to 2, respectively. Overall and intra-familial ACEs were then classified as 0, 1, and 2 or more; and extra-familial ACEs were classified as 0, 1, and 2.

Lastly, the SHARE captured 6 ACEs, including 4 intra-familial domains (emotional neglect, absent biological parent, physical abuse, and economic adversity) and 2 extra-familial domains (non-parental abuse and loneliness)^31^. All domains were summed to obtain overall ACEs, intra-familial ACEs, and extra-familial ACEs with values ranging from 0 to 6, 0 to 4, and 0 to 2, respectively. The ACE classification is the same as which in the HRS.

### Depressive symptoms Assessment

In the CHARLS, depressive symptoms were assessed using the 10-item Center for Epidemiological Studies Depression Scale (CESD-10)^32^, whose validity has been thoroughly demonstrated in the Chinese population^33^. Participants were asked, “how you have felt and behaved during the last week”, with four options of frequency. This study assigned 0-3 to these four options, and the maximum score is 30. The cutoff for depressive symptoms was 10^34,35^.

In the HRS, depressive symptoms were measured by the eight-item Center for Epidemiologic Studies Depression Scale (CESD-8), which has high specificity and sensitivity in identifying depressive cases^36,37^. The CESD-8 contains six negative indicators and two positive indicators. The response “yes” to each negative indicator and “no” to each positive indicator obtained one point. The cutoff for depressive symptoms was 4^38,39^.

In the SHARE, depressive symptoms were assessed using the Europe-depression (EURO-D) scale^40,41^. This scale covers 12 sentiments of the respondent in the previous month, including depressed mood, pessimism, suicidal tendencies, etc. Each item takes 0 or 1 point. The cutoff for depressive symptoms was 4, which has proven to have great consistency and validity^42,43^.

### Covariates

Information on age, sex, race (HRS only), residence (CHARLS and SHARE only), education, economic status, smoking history, and drinking history were self-reported through questionnaires at the baselines of CHARLS, HRS, and SHARE. Race was categorized into White/Caucasian and Black or others. Residence was categorized into rural and urban. In the CHARLS, education was classified as primary school or less, middle school, and high school or above; in the HRS, education was classified as lower than high school, high school, college, and above college; and in the SHARE, education was measured by years spent in education. For economic status, we divided household annual or monthly income from each survey into bottom tertile, middle tertile, and top tertile. Smoking history was classified as never smoking and ever smoking. Drinking history was classified as never drinking and ever drinking in the CHARLS and HRS; in the SHARE, drinking history was categorized into no recent drinking and recent drinking^44^.

In the CHARLS, anthropometric data included body mass index (BMI), waist circumference (WC), and blood pressure. In the HRS, WC, blood pressure, and self-reported BMI were measured. In the SHARE, only BMI were available.

Hypertension was identified if one reported having physician-diagnosed hypertension, and/or taking anti-hypertensive drugs, and/or blood pressure≥ 130/80 mmHg in the CHARLS and HRS. However, in the SHARE, only the first two criteria were used to define hypertension. Diabetes, dyslipidemia (except for HRS), and cardiovascular diseases (CVDs) including stroke and heart diseases were defined by self-reported diagnoses, and/or taking related drugs.

### Statistical analysis

In the three cohorts, the baseline characteristics of included participants were described as medians with interquartile ranges (IQRs) for continuous variables, and frequencies and percentages (%) for categorical variables.

First, frequency tables were used to show the prevalence of spousal concordance of ACEs^13^, with stratification of specific ACE domains. The correlations of each specific ACE between couples were calculated using the Chi-square test or Fisher’s exact test and this generated Cramer’s V. Furthermore, partial Spearman’s correlation test was used to calculate the correlations of overall ACEs, intra-familial ACEs, and extra-familial ACEs between couples.

After excluding couples with missing data on depressive symptoms in the three cohorts, multivariable logistic regression was used to investigate the association of respondents’ ACEs with spousal depressive symptoms. All estimates were adjusted for spousal ACEs, age, race (HRS only), residence (CHARLS and SHARE only), education, economic status, BMI, WC (CHARLS and HRS only), smoking history, drinking history, diabetes, hypertension, dyslipidemia (CHARLS and SHARE only), and CVDs. We also assessed the association between respondents’ ACEs and depressive symptoms of their spouses who had never experienced ACEs.

Finally, we performed mediation analyses to explore whether respondents’ depressive symptoms mediated the effect of respondents’ ACEs on spousal depressive symptoms. Reporting of this study was done under Strengthening the Reporting of Observational studies in Epidemiology (STROBE) guidelines^45^. Analyses were performed using SAS statistical software version 9.4 (SAS Institute) and R statistical software version 4.1.2 (R Project for Statistical Computing). All analyses were two-sided, and a *P* value of < 0.05, a 95% confidence interval (CI) of odds ratio (OR) that did not cross 1.00, or a 95% CI of Spearman’s correlation or β that did not cross 0 was considered statistically significant.

## Results

### Population characteristics

The baseline characteristics of participant couples from the CHARLS, HRS, and SHARE are shown in **eTable 2**. Among 2394 couples in the CHARLS, more than 90% of these couples experienced ACEs. More specifically, intra-familial ACEs affected 92.2% of husbands and 89.1% of wives, while 29.5% of husbands and 27.2% of wives reported having experienced extra-familial ACEs. Among 3131 couples from the HRS, 60.4% of husbands and 52.2% of wives reported having ACEs. The proportions of husbands exposed to intra-familial ACEs and extra-familial ACEs were 48.4% and 29.0%, while of wives 48.4% and 11.8%. Among 1831 spouses in the SHARE, about 60% suffered from ACEs. Intra-familial ACEs affected 53.8% of husbands and 50% of wives, while extra-familial ACEs impacted29.7% of husbands and 23.1% of wives.

### Spousal concordance of ACEs

The majority (more than 50%) of couples from the three cohorts were in concordance across specific and multiple domains of ACEs (**Table 1**). In the CHARLS, domain-specific analyses indicated significant correlations between couples except for parental separation or divorce, parents incarcerated, and sibling death. In the HRS, significant correlations of all specific ACEs were found between couples. Among those in the SHARE, noticeable correlations were found except for absent biological parents. After adjusting for both couples’ characteristics, overall ACEs, intra-familial ACEs, and extra-familial ACEs showed substantial correlations between spouses in the CHARLS, HRS, and SHARE (**eTable 3**).

**Table 1.** Spousal concordance of domains of ACEs in the CHARLS, HRS, and SHARE

| Domains of ACEs | Concordant (%) |  | Discordant: 1 spouse had (%) | Cramer's V | P value |
| --- | --- | --- | --- | --- | --- |
|  | Both had | Both did not have |  |  |  |
| *CHARLS* |  |  |  |  |  |
| *Specific domains* |  |  |  |  |  |
| Emotional neglect | 19.0 | 39.8 | 41.2 | 0.14 | <0.001 |
| Family violence | 7.9 | 60.1 | 32.1 | 0.12 | <0.001 |
| Parental separation or divorce | 0.0 | 99.0 | 1.0 | / | 1.000 |
| Parental substance abuse | 37.2 | 25.1 | 37.7 | 0.24 | <0.001 |
| Parents incarcerated | 0.0 | 99.5 | 0.5 | / | 1.000 |
| Parental mental illness | 2.9 | 80.2 | 16.9 | 0.16 | <0.001 |
| Parental disability | 5.9 | 64.1 | 30.0 | 0.09 | <0.001 |
| Parental death | 2.2 | 76.2 | 21.6 | 0.05 | 0.029 |
| Sibling death | 0.3 | 90.6 | 9.1 | 0.02 | 0.489 |
| Physical abuse | 11.9 | 51.9 | 36.2 | 0.15 | <0.001 |
| Economic adversity | 16.4 | 44.6 | 39.1 | 0.15 | <0.001 |
| Bullying | 4.3 | 71.1 | 24.6 | 0.12 | <0.001 |
| Loneliness | 2.0 | 81.9 | 16.1 | 0.11 | <0.001 |
| Community violence | 2.4 | 82.2 | 15.5 | 0.15 | <0.001 |
| *Overall ACEs* |  |  |  | 0.20 | <0.001 |
| 1 or more | 85.5 | 1.4 | 13.1 |  |  |
| 2 or more | 54.9 | 10.2 | 34.9 |  |  |
| 3 or more | 28.0 | 28.5 | 43.5 |  |  |
| 4 or more | 11.5 | 51.7 | 36.8 |  |  |
| *Intra-familial ACEs* |  |  |  | 0.17 | <0.001 |
|  | Both had | Both did not have |  |  |  |
| 1 or more | 83.2 | 2.0 | 14.8 |  |  |
| 2 or more | 49.4 | 13.2 | 37.5 |  |  |
| 3 or more | 20.2 | 35.2 | 44.6 |  |  |
| 4 or more | 6.4 | 62.5 | 31.2 |  |  |
| *Extra-familial ACEs* |  |  |  | 0.17 | <0.001 |
| 1 or more | 11.5 | 54.8 | 33.7 |  |  |
| 2 or more | 0.8 | 86.9 | 12.3 |  |  |
| **HRS** |  |  |  |  |  |
| *Specific domains* |  |  |  |  |  |
| Emotional neglect | 3.3 | 70.7 | 26.0 | 0.05 | 0.010 |
| Parental substance abuse | 4.5 | 68.1 | 27.4 | 0.08 | <0.001 |
| Physical abuse | 0.9 | 86.2 | 12.8 | 0.06 | 0.002 |
| Economic adversity | 10.2 | 53.7 | 36.1 | 0.11 | <0.001 |
| Repeating school year | 3.6 | 72.6 | 23.8 | 0.12 | <0.001 |
| Trouble with police | 0.6 | 86.4 | 13.0 | 0.09 | <0.001 |
| *Overall ACEs* |  |  |  | 0.14 | <0.001 |
| 1 or more | 34.3 | 21.7 | 44.0 |  |  |
| 2 or more | 7.7 | 59.4 | 32.9 |  |  |
| *Intra-familial ACEs* |  |  |  | 0.10 | <0.001 |
| 1 or more | 25.8 | 29.0 | 45.2 |  |  |
| 2 or more | 3.7 | 71.3 | 25.0 |  |  |
| *Extra-familial ACEs* |  |  |  | 0.10 | <0.001 |
| 1 or more | 4.8 | 63.9 | 31.3 |  |  |
| 2 | 0.2 | 95.8 | 4.1 |  |  |
|  | Both had | Both did not have |  |  |  |
| *SHARE* |  |  |  |  |  |
| *Specific domains* |  |  |  |  |  |
| Emotional neglect | 8.1 | 63.9 | 28.0 | 0.19 | <0.001 |
| Absent biological parent | 1.9 | 78.4 | 19.8 | 0.05 | 0.055 |
| Physical abuse | 7.7 | 66.9 | 25.5 | 0.23 | <0.001 |
| Economic adversity | 11.6 | 63.8 | 24.6 | 0.33 | <0.001 |
| Non-parental abuse | 1.0 | 91.3 | 7.7 | 0.17 | <0.001 |
| Loneliness | 7.4 | 70.5 | 22.1 | 0.27 | <0.001 |
| *Overall ACEs* |  |  |  | 0.29 | <0.001 |
| 1 or more | 41.8 | 22.4 | 35.8 |  |  |
| 2 or more | 11.3 | 56.7 | 32.0 |  |  |
| *Intra-familial ACEs* |  |  |  | 0.24 | <0.001 |
| 1 or more | 32.3 | 28.5 | 39.3 |  |  |
| 2 or more | 6.7 | 66.6 | 26.8 |  |  |
| *Extra-familial ACEs* |  |  |  | 0.27 | <0.001 |
| 1 or more | 8.9 | 66.1 | 25.0 |  |  |
| 2 | 0.2 | 96.3 | 3.6 |  |  |

Abbreviations: ACEs, adverse childhood experiences. CHARLS, China Health and Retirement Longitudinal Study. HRS, Health and Retirement Study. SHARE, Survey of Health, Ageing and Retirement in Europe.

### Association of respondents’ ACEs with spousal depressive symptoms

Overall, 2263 husbands (24.2% cases of depressive symptoms) and 2203 wives (38.0% cases of depressive symptoms) were included in the CHARLS. We found significant associations between husbands’ ACEs and wives’ depressive symptoms, with fully adjusted ORs (95% CI) of 1.61 (1.03-2.50), 2.09 (1.36-3.22), and 1.13 (1.07-1.19) for 2, 4 or more, and continuous ACEs, respectively. On the other hand, wives with ACEs (continuous) were associated with a significantly higher risk of husbands’ depressive symptoms (OR=1.07, 95% CI 1.02 to 1.13), as shown in **Table 2**. More information is provided in **eTable 4**.

**Table 2.**
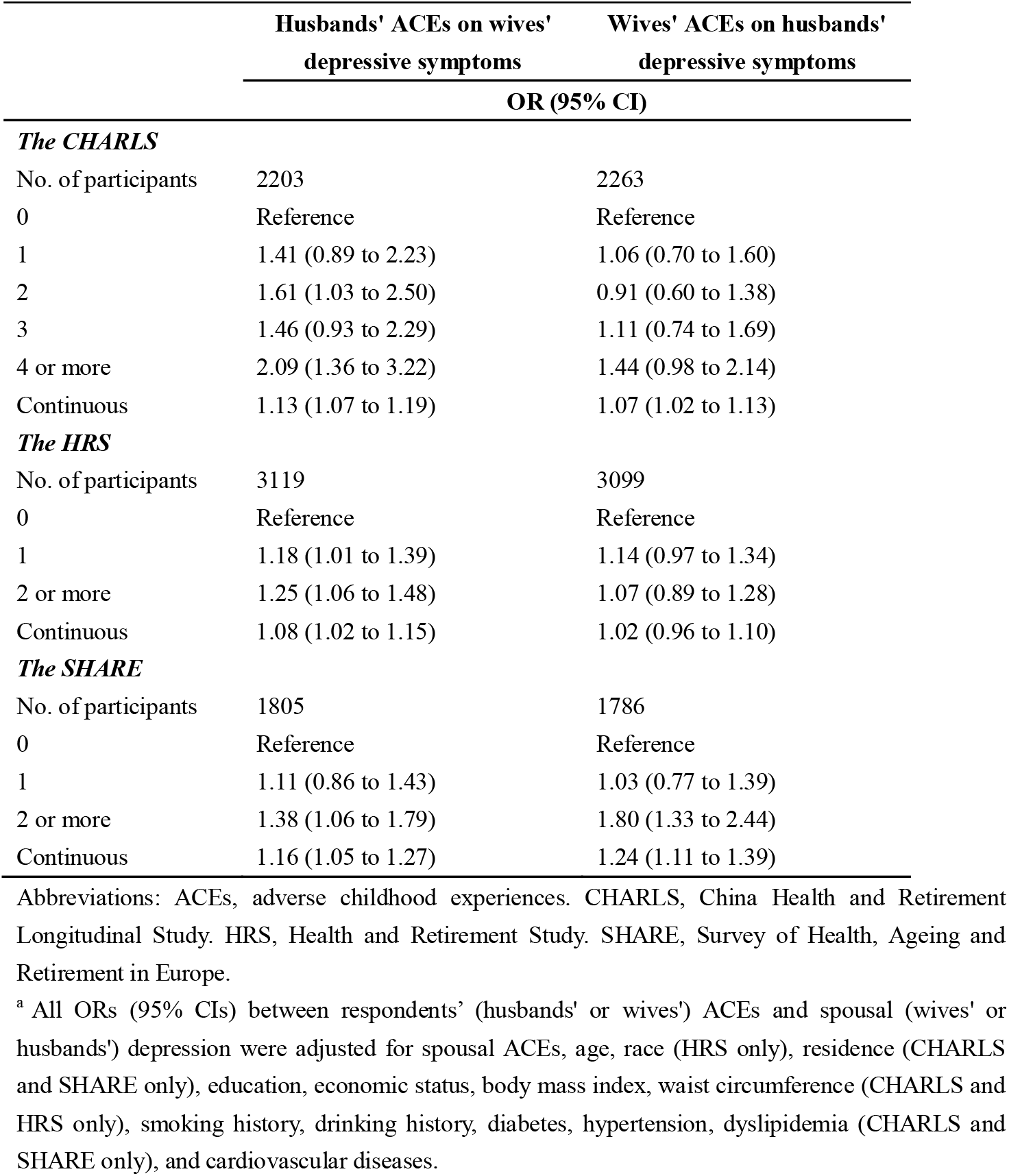
Association between respondents’ overall ACEs and spousal depressive symptoms in the CHARLS, HRS, and SHARE^a^

|  | Husbands' ACEs on wives'<br>depressive symptoms | Wives' ACEs on husbands'<br>depressive symptoms |
| --- | --- | --- |
|  | OR (95% CI) |  |
| *The CHARLS* |  |  |
| No. of participants | 2203 | 2263 |
| 0 | Reference | Reference |
| 1 | 1.41 (0.89 to 2.23) | 1.06 (0.70 to 1.60) |
| 2 | 1.61 (1.03 to 2.50) | 0.91 (0.60 to 1.38) |
| 3 | 1.46 (0.93 to 2.29) | 1.11 (0.74 to 1.69) |
| 4 or more | 2.09 (1.36 to 3.22) | 1.44 (0.98 to 2.14) |
| Continuous | 1.13 (1.07 to 1.19) | 1.07 (1.02 to 1.13) |
| *The HRS* |  |  |
| No. of participants | 3119 | 3099 |
| 0 | Reference | Reference |
| 1 | 1.18 (1.01 to 1.39) | 1.14 (0.97 to 1.34) |
| 2 or more | 1.25 (1.06 to 1.48) | 1.07 (0.89 to 1.28) |
| Continuous | 1.08 (1.02 to 1.15) | 1.02 (0.96 to 1.10) |
| *The SHARE* |  |  |
| No. of participants | 1805 | 1786 |
| 0 | Reference | Reference |
| 1 | 1.11 (0.86 to 1.43) | 1.03 (0.77 to 1.39) |
| 2 or more | 1.38 (1.06 to 1.79) | 1.80 (1.33 to 2.44) |
| Continuous | 1.16 (1.05 to 1.27) | 1.24 (1.11 to 1.39) |

Abbreviations: ACEs, adverse childhood experiences. CHARLS, China Health and Retirement Longitudinal Study. HRS, Health and Retirement Study. SHARE, Survey of Health, Ageing and Retirement in Europe.
<sup>a</sup> All ORs (95% CIs) between respondents' (husbands' or wives') ACEs and spousal (wives' or husbands') depression were adjusted for spousal ACEs, age, race (HRS only), residence (CHARLS and SHARE only), education, economic status, body mass index, waist circumference (CHARLS and HRS only), smoking history, drinking history, diabetes, hypertension, dyslipidemia (CHARLS and SHARE only), and cardiovascular diseases.

In the HRS, a total of 3099 husbands and 3119 wives were included, of whom around 10% had depressive symptoms. Wives whose husbands had 2 or more ACEs were 25% more likely to have depressive symptoms compared to those whose spouses had no ACE. However, wives’ ACEs were not significantly associated with husbands’ depressive symptoms.

In the SHARE, the proportion of depressive symptoms among the included 1786 husbands and 1850 wives were 21.4% and 32.7%, respectively. It was found that husbands’ 2 or more ACEs (vs. 0 ACE) were significantly associated with wives’ depressive symptoms, with fully adjusted ORs (95% CIs) of 1.38 (1.06 to 1.79). Similarly, wives’ ACEs were associated with husbands’ depressive symptoms, with ORs (95% CIs) of 1.80 (1.33 to 2.44) for 2 or more ACEs as shown in **Table 2**.

Furthermore, the associations between respondents’ ACEs and the depressive symptoms of spouses who had no ACEs are shown in **Table 3**. In the CHARLS, we did not observe significant associations between respondents’ ACEs and spousal depressive symptoms. In the HRS, the ORs (95% CIs) of husbands’ 1 ACE and 2 or more ACEs (vs. 0 ACE) on wives’ depressive symptoms were 1.32 (1.05 to 1.66) and 1.40 (1.08 to 1.81), respectively. Wives’ 1 ACE was also found to be associated with husbands’ depressive symptoms. In the SHARE, a substantial association was only found between wives’ 2 or more ACEs (vs. 0 ACE) and husbands’ depressive symptoms (OR=2.04, 95% CI 1.15 to 3.61).

**Table 3.** Association between respondents’ ACEs and the depressive symptoms of spouses who had no ACEs in the CHARLS, HRS, and SHARE^a^

|  | Husbands' ACEs on wives'<br>depressive symptoms | Wives' ACEs on husbands'<br>depressive symptoms |
| --- | --- | --- |
|  | OR (95% CI) |  |
| *The CHARLS* |  |  |
| No. of participants | 211 | 152 |
| 0 | Reference | Reference |
| 1 | 1.47 (0.47 to 4.58) | 2.79 (0.62 to 12.50) |
| 2 or more | 1.22 (0.45 to 3.33) | 1.15 (0.27 to 4.99) |
| Continuous | 1.06 (0.88 to 1.27) | 0.84 (0.60 to 1.17) |
| *The HRS* |  |  |
| No. of participants | 1495 | 1239 |
| 0 | Reference | Reference |
| 1 | 1.32 (1.05 to 1.66) | 1.45 (1.11 to 1.89) |
| 2 or more | 1.40 (1.08 to 1.81) | 1.23 (0.89 to 1.71) |
| Continuous | 1.14 (1.04 to 1.26) | 1.12 (0.99 to 1.27) |
| *The SHARE* |  |  |
| No. of participants | 733 | 710 |
| 0 | Reference | Reference |
| 1 | 0.93 (0.61 to 1.42) | 0.93 (0.56 to 1.55) |
| 2 or more | 1.14 (0.72 to 1.81) | 2.04 (1.15 to 3.61) |
| Continuous | 1.05 (0.88 to 1.26) | 1.23 (0.99 to 1.55) |

Abbreviations: ACEs, adverse childhood experiences. CHARLS, China Health and Retirement Longitudinal Study. HRS, Health and Retirement Study. SHARE, Survey of Health, Ageing and Retirement in Europe.
<sup>a</sup> All ORs (95% CIs) between respondents' (husbands' or wives') ACEs and spousal (wives' or husbands') depression were adjusted for spousal age, race (HRS only), residence (CHARLS and SHARE only), education, economic status, body mass index, waist circumference (CHARLS and HRS only), smoking history, drinking history, diabetes, hypertension, dyslipidemia (CHARLS and SHARE only), and cardiovascular diseases.

### Mediation of respondents’ ACEs and spousal depressive symptoms by respondents’ depressive symptoms

Finally, we found that respondents’ depressive symptoms mediated the effect of respondents’ ACEs on spousal depressive symptoms, as shown in **Table 4**. More specifically, husbands’ depressive symptoms mediated 23.5% (5.2% to 52.0%), 24.2% (2.7% to 129.5%), and 29.4% (4.5% to 79.2%) of the effect of husbands’ ACEs on wives’ depressive symptoms in the CHARLS, HRS, and SHARE, respectively. Furthermore, wives’ depressive symptoms mediated 67.9% (23.1% to 270.9%) and 36.9% (14.7% to 71.7%) of the effect of wives’ ACEs on husbands’ depressive symptoms in the CHARLS and SHARE, respectively.

**Table 4.**
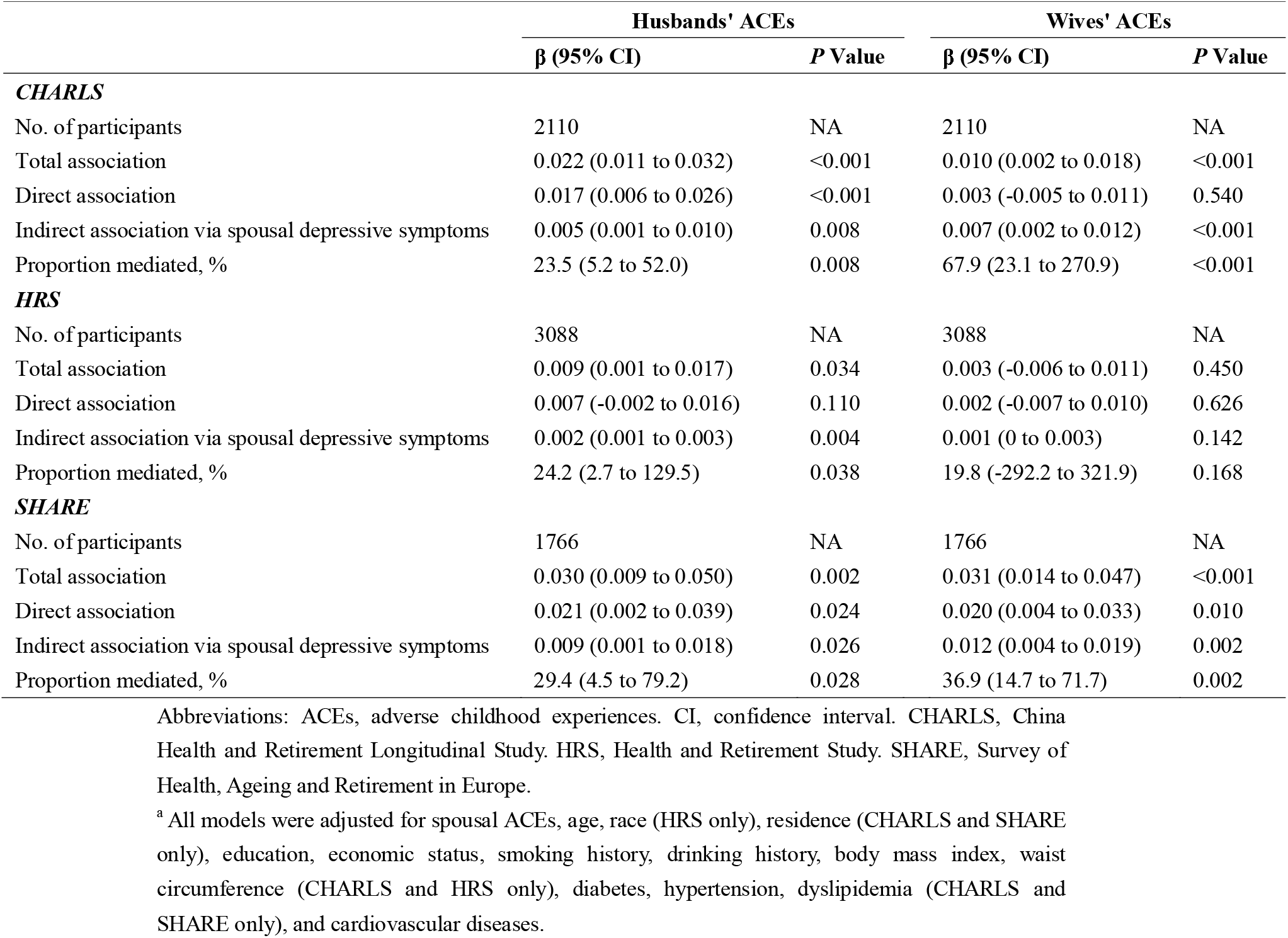
Adjusted direct and indirect associations of respondents’ ACEs with spousal depressive symptoms via respondents’ depression^a^

## Discussion

Across the CHARLS, HRS, and SHARE, there was considerable concordance of overall ACEs, intra-familial ACEs, and extra-familial ACEs between spouses. Except for wives’ ACEs and husbands’ depressive symptoms in the HRS, respondents’ ACEs, especially the intra-familial ACEs, were significantly associated with higher spousal depressive risks in all three cohorts, with more than 20% of these associations being mediated by respondents’ depressive symptoms.

Spousal concordance in ACEs can be explained by the assortative mating theory, which proposes that people are inclined to marry partners who are similar to them in some way, such as socioeconomic status, lifestyle, and even cardiovascular risk^13,46,47^. A recent study also found that women who experienced ACEs tended to have husbands with ACEs as well^48^. Moreover, ACEs themselves can increase the risks of lower socioeconomic status, substance abuse, and cardiovascular risks, then those consequences may further influence mate choice in different ways^8,49^.

We also found significant associations between respondents’ ACEs and spousal depressive symptoms in middle-aged and older adults even after adjusting for spousal ACEs, and these associations were mediated by respondents’ depressive symptoms. It is well known that individuals with ACEs are prone to developing depression^5,50^.

Previous studies have revealed that spouses of those who suffer from depression experience high marital distress and low marital satisfaction, which may further exacerbate their depressive symptoms^51,52^. In addition, respondents’ depressive symptoms can independently aggravate spousal depressive symptoms^53^. Physically, ACEs can exacerbate the hypothalamic-pituitary-adrenal (HPA) axis dysfunction by inducing couple conflict, resulting in reduced abilities of vital organs to respond to environmental stimulation like stress and emotion, leading to depressive symptoms in spouses^54-57^. From the perspective of the couple-based relationship, the stress of one person has marked impacts on other intimate members in the household, which is called stress contagion within couples^16,58^. Relationship distress exists because couples frequently interact in maladaptive ways around depressive symptoms, whether one or both partners suffer from them. Due to the interdependence of spousal lives, the presence of relationship distress predicts the development of depressive symptoms in one partner, and this increases the possibility of the other spouse becoming distressed and obtaining depressive symptoms^59^.

Interestingly, this study found that the association of husbands’ ACEs with wives’ depressive symptoms was more significant than vice versa. Prior research has shown that wives respond more than husbands to their spouses’ chronic health conditions, and this aligns with the results from this study^60^. Furthermore, when couples experienced negative communications, wives tended to have worse sleep quality than husbands^61^. Poorer sleep can further stimulate the production of pro-inflammatory cytokines and promote depressive symptoms^62^. Given that women are more affected by sleep disorders and inflammation than men^63^, ACEs may induce increased marital conflict and render wives more susceptible to depressive symptoms.

To the best of our knowledge, this is the first and most comprehensive study to explore the spousal concordance of ACEs, the association between respondents’ ACEs and spousal depressive symptoms, and the mediating role of respondents’ depressive symptoms in this association. We used data from three rigorous and valid surveys, the CHARLS, HRS, and SHARE from China, the US, and Europe respectively. These three regions represent three different levels of development in terms of public welfare. The results of the study showed that across these populations, ACEs are significantly correlated between couples, and respondents’ depressive symptoms mediated more than 20% of the effect of respondents’ spousal ACEs on depressive symptoms. In addition, this study divided overall ACEs into intra-familial and extra-familial ACEs to explore their spousal concordance and associations with spousal depression, filling an important gap in the literature.

There are still some limitations. First, information on ACEs was collected via self-reported questionnaires, which may have led to recall bias. Second, the specific items of ACEs and definitions of several covariates differ across the three cohorts because of different questionnaire designs. Finally, due to the retrospective design, the sequence in which couples experience depressive symptoms is unclear.

## Conclusion

We found that couples’ overall, intra-familial, and extra-familial ACEs were significantly correlated. Additionally, respondents’ ACEs were associated with spousal depressive symptoms with respondents’ depressive symptoms as a mediator, which urges us to consider the implications of childhood adversity on later-life depressive symptoms at the couple-level. It is necessary to strengthen the effective interventions of ACEs and depressive symptoms to reduce the detrimental influence on individuals and the two-way influence between couples.

## Data Availability

All data produced in the present work are contained in the manuscript

## Authors’ contributions

PS and ZR designed the study. ZR managed and analyzed the data. ZR and WS prepared the first draft. ZR, WS, SZ, SC and WL reviewed and edited the manuscript, with comments from PS, HCW L, WX, CY, DA, IR, DC. All authors were involved in revising the paper, had full access to the data, and gave final approval of the submitted versions.

## Data sharing statement

The data that support the findings of this study are available from the websites of China Health and Retirement Longitudinal Study at http://charls.pku.edu.cn/en, Health and Retirement Study at https://hrsdata.isr.umich.edu/, and Survey of Health, Ageing and Retirement in Europe at http://www.share-project.org/home0.html. The SAS and R code for this study can be requested from the corresponding author (Prof. PS).

## Acknowledgments

We are grateful to the China Center for Economic Research at Beijing University for providing us with the data, and we thank the CHARLS research and field team for collecting the data.

The HRS (Health and Retirement Study) is sponsored by the National Institute on Aging (grant number NIA U01AG009740) and is conducted by the University of Michigan.

The SHARE data collection has been funded by the European Commission, DG RTD through FP5 (QLK6-CT-2001-00360), FP6 (SHARE-I3: RII-CT-2006-062193, COMPARE: CIT5-CT-2005-028857, SHARELIFE: CIT4-CT-2006-028812), FP7 (SHARE-PREP: GA N°211909, SHARE-LEAP: GA N°227822, SHARE M4: GA N°261982, DASISH: GA N°283646) and Horizon 2020 (SHARE-DEV3: GA N°676536, SHARE-COHESION: GA N°870628, SERISS: GA N°654221, SSHOC: GA N°823782, SHARE-COVID19: GA N°101015924) and by DG Employment, Social Affairs & Inclusion through VS 2015/0195, VS 2016/0135, VS 2018/0285, VS 2019/0332, and VS 2020/0313. Additional funding from the German Ministry of Education and Research, the Max Planck Society for the Advancement of Science, the U.S. National Institute on Aging (U01_AG09740-13S2, P01_AG005842, P01_AG08291, P30_AG12815, R21_AG025169, Y1-AG-4553-01, IAG_BSR06-11, OGHA_04-064, HHSN271201300071C, RAG052527A) and from various national funding sources is gratefully acknowledged (see www.share-project.org).

## Reference

1. McCarron RM, Shapiro B, Rawles J, Luo J. Depression. Ann Intern Med. 2021;174(5):Itc65–itc80.

2. Global, regional, and national burden of 12 mental disorders in 204 countries and territories, 1990-2019: a systematic analysis for the Global Burden of Disease Study 2019. Lancet Psychiatry. 2022;9(2):137–150.

3. Malhi GS, Mann JJ. Depression. Lancet. 2018;392(10161):2299–2312.

4. König H, König HH, Konnopka A. The excess costs of depression: a systematic review and meta-analysis. Epidemiol Psychiatr Sci. 2019;29:e30.

5. Hughes K, Bellis MA, Hardcastle KA, et al. The effect of multiple adverse childhood experiences on health: a systematic review and meta-analysis. Lancet Public health. 2017;2(8):e356–e366.

6. Masuda A, Yamanaka T, Hirakawa T, et al. Intra- and extra-familial adverse childhood experiences and a history of childhood psychosomatic disorders among Japanese university students. BioPsychoSocial medicine. 2007;1:9.

7. Nelson CA, Scott RD, Bhutta ZA, Harris NB, Danese A, Samara M. Adversity in childhood is linked to mental and physical health throughout life. BMJ. 2020;371:m3048.

8. Bellis MA, Hughes K, Ford K, Ramos Rodriguez G, Sethi D, Passmore J. Life course health consequences and associated annual costs of adverse childhood experiences across Europe and North America: a systematic review and meta-analysis. Lancet Public health. 2019;4(10):e517–e528.

9. Hughes K, Ford K, Bellis MA, Glendinning F, Harrison E, Passmore J. Health and financial costs of adverse childhood experiences in 28 European countries: a systematic review and meta-analysis. Lancet Public health. 2021;6(11):e848–e857.

10. Liu M, Luong L, Lachaud J, Edalati H, Reeves A, Hwang SW. Adverse childhood experiences and related outcomes among adults experiencing homelessness: a systematic review and meta-analysis. Lancet Public health. 2021;6(11):e836–e847.

11. Pradeep N, Sutin AR. Spouses and depressive symptoms in older adulthood. Sci Rep. 2015;5:8594.

12. Homish GG, Leonard KE, Kearns-Bodkin JN. Alcohol use, alcohol problems, and depressive symptomatology among newly married couples. Drug Alcohol Depend. 2006;83(3):185–192.

13. Retnakaran R, Wen SW, Tan H, et al. Spousal Concordance of Cardiovascular Risk Factors in Newly Married Couples in China. JAMA Netw Open. 2021;4(12):e2140578.

14. Felitti VJ, Anda RF, Nordenberg D, et al. REPRINT OF: Relationship of Childhood Abuse and Household Dysfunction to Many of the Leading Causes of Death in Adults: The Adverse Childhood Experiences (ACE) Study. Am J Prev Med. 2019;56(6):774–786.

15. Sonu S, Post S, Feinglass J. Adverse childhood experiences and the onset of chronic disease in young adulthood. Prev Med. 2019;123:163–170.

16. Lam J. Actor-Partner Effects of Childhood Disadvantage on Later Life Subjective Well-Being Among Individuals in Coresidential Unions. J Gerontol B Psychol Sci Soc Sci. 2020;75(6):1275–1285.

17. Zhao Y, Hu Y, Smith JP, Strauss J, Yang G. Cohort profile: the China Health and Retirement Longitudinal Study (CHARLS). Int J Epidemiol. 2014;43(1):61–68.

18. Sonnega A, Faul JD, Ofstedal MB, Langa KM, Phillips JW, Weir DR. Cohort Profile: the Health and Retirement Study (HRS). Int J Epidemiol. 2014;43(2):576–585.

19. Börsch-Supan A, Brandt M, Hunkler C, et al. Data Resource Profile: the Survey of Health, Ageing and Retirement in Europe (SHARE). Int J Epidemiol. 2013;42(4):992–1001.

20. Börsch-Supan A. Survey of Health, Ageing and Retirement in Europe (SHARE) Wave 8. Release version: 8.0.0. SHARE-ERIC. Data set. DOI: 10.6103/SHARE.w8.800.

21. Bergmann MaAB-SE. SHARE Wave 8 Methodology: Collecting Cross-National Survey Data in Times of COVID-19. Munich: MEA, Max Planck Institute for Social Law and Social Policy.

22. Börsch-Supan A. Survey of Health, Ageing and Retirement in Europe (SHARE) Wave 7. Release version: 8.0.0. SHARE-ERIC. Data set. DOI: 10.6103/SHARE.w7.800.

23. Bergmann M, A. Scherpenzeel and A. Börsch-Supan (Eds.) (2019). SHARE Wave 7 Methodology: Panel Innovations and Life Histories. Munich: MEA, Max Planck Institute for Social Law and Social Policy.

24. Giovanelli A, Reynolds AJ. Adverse childhood experiences in a low-income black cohort: The importance of context. Prev Med. 2021;148:106557.

25. Lin L, Wang HH, Lu C, Chen W, Guo VY. Adverse Childhood Experiences and Subsequent Chronic Diseases Among Middle-aged or Older Adults in China and Associations With Demographic and Socioeconomic Characteristics. JAMA Netw Open. 2021;4(10):e2130143.

26. Schneider S. Associations between childhood exposure to community violence, child maltreatment and school outcomes. Child Abuse Negl. 2020;104:104473.

27. Xerxa Y, Rescorla LA, Shanahan L, Tiemeier H, Copeland WE. Childhood loneliness as a specific risk factor for adult psychiatric disorders. Psychol Med. 2021:1–9.

28. Klomek AB, Sourander A, Elonheimo H. Bullying by peers in childhood and effects on psychopathology, suicidality, and criminality in adulthood. Lancet Psychiatry. 2015;2(10):930–941.

29. Xiang X, Wang X. Childhood adversity and major depression in later life: A competing-risks regression analysis. Int J Geriatr Psychiatry. 2021;36(1):215–223.

30. Krause N, Shaw BA, Cairney J. A descriptive epidemiology of lifetime trauma and the physical health status of older adults. Psychol Aging. 2004;19(4):637–648.

31. Brugiavini A, Buia R, Kovacic M, Orso Cjssep. Adverse Childhood Experiences and Outcomes Later in Life: Evidence from SHARE Countries.

32. Andresen EM, Malmgren JA, Carter WB, Patrick DL. Screening for depression in well older adults: evaluation of a short form of the CES-D (Center for Epidemiologic Studies Depression Scale). Am J Prev Med. 1994;10(2):77–84.

33. Chen H, Mui AC. Factorial validity of the Center for Epidemiologic Studies Depression Scale short form in older population in China. Int Psychogeriatr. 2014;26(1):49–57.

34. Zhou L, Ma X, Wang W. Relationship between Cognitive Performance and Depressive Symptoms in Chinese Older Adults: The China Health and Retirement Longitudinal Study (CHARLS). J Affect Disord. 2021;281:454–458.

35. Li H, Liu X, Zheng Q, Zeng S, Luo X. Gender differences and determinants of late-life depression in China: A cross-sectional study based on CHARLS. J Affect Disord. 2022;309:178–185.

36. Zivin K, Pirraglia PA, McCammon RJ, Langa KM, Vijan S. Trends in depressive symptom burden among older adults in the United States from 1998 to 2008. J Gen Intern Med. 2013;28(12):1611–1619.

37. Dang L, Dong L, Mezuk B. Shades of Blue and Gray: A Comparison of the Center for Epidemiologic Studies Depression Scale and the Composite International Diagnostic Interview for Assessment of Depression Syndrome in Later Life. Gerontologist. 2020;60(4):e242–e253.

38. Gould CE, Rideaux T, Spira AP, Beaudreau SA. Depression and anxiety symptoms in male veterans and non-veterans: the Health and Retirement Study. Int J Geriatr Psychiatry. 2015;30(6):623–630.

39. Quiñones AR, Markwardt S, Botoseneanu A. Diabetes-Multimorbidity Combinations and Disability Among Middle-aged and Older Adults. J Gen Intern Med. 2019;34(6):944–951.

40. Han FF, Wang HX, Wu JJ, Yao W, Hao CF, Pei JJ. Depressive symptoms and cognitive impairment: A 10-year follow-up study from the Survey of Health, Ageing and Retirement in Europe. Eur Psychiatry. 2021;64(1):e55.

41. Wu JJ, Wang HX, Yao W, Yan Z, Pei JJ. Late-life depression and the risk of dementia in 14 countries: a 10-year follow-up study from the Survey of Health, Ageing and Retirement in Europe. J Affect Disord. 2020;274:671–677.

42. Guerra M, Ferri C, Llibre J, Prina AM, Prince M. Psychometric properties of EURO-D, a geriatric depression scale: a cross-cultural validation study. BMC Psychiatry. 2015;15:12.

43. Prince MJ, Reischies F, Beekman ATF, et al. Development of the EURO–D scale – a European Union initiative to compare symptoms of depression in 14 European centres. Br J Psychiatry. 1999;174(4):330–338.

44. Hajek A, König HH. Asymmetric effects of obesity on loneliness among older Germans. Longitudinal findings from the Survey of Health, Ageing and Retirement in Europe. Aging Ment Health. 2021;25(12):2293–2297.

45. von Elm E, Altman DG, Egger M, Pocock SJ, Gøtzsche PC, Vandenbroucke JP. Strengthening the Reporting of Observational Studies in Epidemiology (STROBE) statement: guidelines for reporting observational studies. BMJ. 2007;335(7624):806–808.

46. Howe LJ, Lawson DJ, Davies NM, et al. Genetic evidence for assortative mating on alcohol consumption in the UK Biobank. Nat Commun. 2019;10(1):5039.

47. Zhang L, Tan X. Educational Assortative Mating and Health: A Study in Chinese Internal Migrants. Int J Environ Res Public Health. 2021;18(4).

48. Andersson SO, Annerbäck EM, Söndergaard HP, Hallqvist J, Kristiansson P. Adverse Childhood Experiences are associated with choice of partner, both partners’ relationship and psychosocial health as reported one year after birth of a common child. A cross-sectional study. PLoS One. 2021;16(1):e0244696.

49. Houtepen LC, Heron J, Suderman MJ, Fraser A, Chittleborough CR, Howe LD. Associations of adverse childhood experiences with educational attainment and adolescent health and the role of family and socioeconomic factors: A prospective cohort study in the UK. PLoS Med. 2020;17(3):e1003031.

50. Salas J, van den Berk-Clark C, Skiöld-Hanlin S, Schneider FD, Scherrer JF. Adverse childhood experiences, depression, and cardiometabolic disease in a nationally representative sample. J Psychosom Res. 2019;127:109842.

51. Aggarwal S, Kataria D, Prasad S. A comparative study of quality of life and marital satisfaction in patients with depression and their spouses. Asian J Psychiatr. 2017;30:65–70.

52. Whisman MA, Sbarra DA, Beach SRH. Intimate Relationships and Depression: Searching for Causation in the Sea of Association. Annu Rev Clin Psychol. 2021;17:233–258.

53. Kivelä SL, Luukinen H, Viramo P, Koski K. Depression in elderly spouse pairs. Int Psychogeriatr. 1998;10(3):329–338.

54. Holsboer F. The corticosteroid receptor hypothesis of depression. Neuropsychopharmacology. 2000;23(5):477–501.

55. Arbel R, Rodriguez AJ, Margolin G. Cortisol Reactions During Family Conflict Discussions: Influences of Wives’ and Husbands’ Exposure to Family-of-Origin Aggression. Psychol Violence. 2016;6(4):519–528.

56. Rodriguez AJ, Margolin G. Wives’ and husbands’ cortisol reactivity to proximal and distal dimensions of couple conflict. Fam Process. 2013;52(3):555–569.

57. Oyola MG, Handa RJ. Hypothalamic-pituitary-adrenal and hypothalamic-pituitary-gonadal axes: sex differences in regulation of stress responsivity. Stress. 2017;20(5):476–494.

58. Settersten Jr RAJRiHD. Relationships in time and the life course: The significance of linked lives. 2015;12(3-4):217–223.

59. Baucom DH, Fischer MS, Worrell M, et al. Couple-based Intervention for Depression: An Effectiveness Study in the National Health Service in England. Fam Process. 2018;57(2):275–292.

60. Berg CA, Upchurch R. A developmental-contextual model of couples coping with chronic illness across the adult life span. Psychol Bull. 2007;133(6):920–954.

61. Kiecolt-Glaser JK, Wilson SJ. Lovesick: How Couples’ Relationships Influence Health. Annu Rev Clin Psychol. 2017;13:421–443.

62. Irwin MR, Olmstead RE, Ganz PA, Haque R. Sleep disturbance, inflammation and depression risk in cancer survivors. Brain Behav Immun. 2013;30 Suppl(Suppl)S58–67.

63. Dolsen MR, Crosswell AD, Prather AA. Links Between Stress, Sleep, and Inflammation: Are there Sex Differences? Curr Psychiatry Rep. 2019;21(2):8.

