## Supplement for "Spousal concordance in adverse childhood experiences and the association with depressive symptoms in middle-aged and older adults: findings across China, the US, and Europe"

Manuscript word count: 2992

**eTable 1.** Definition of ACEs in the CHARLS, HRS, and SHARE

**eTable 2.** Baseline Characteristics of included couples in the CHARLS, HRS, and SHARE

**eTable 3.** Adjusted Spearman correlations of spousal multiple ACEs in the CHARLS, HRS, and SHARE

**eTable 4.** Association of respondents’ intra-familial and extra-familial ACEs with spousal depressive symptoms in the CHARLS, HRS, and SHARE

**eFigure 1.** Flow chart of analyses in the CHARLS

**eFigure 2.** Flow chart of analyses in the HRS

**eFigure 3.** Flow chart of analyses in the SHARE

**eTable 1. Definition of ACEs in the CHARLS, HRS, and SHARE**

| **Domains of ACEs** | | | **Questionnaire Items** | **Answers defined as exposure to the domain** |
| --- | --- | --- | --- | --- |
| CHARLS | Intra-familial | Emotional neglect | How much love and affection did your female guardian give you while you were growing up? | rarely/never |
|  |  |  | How much effort did your female guardian put into watching over you? | a little/not at all |
|  |  |  | Did your male/female guardian treat your siblings better than you when you were growing up? | very strict/somewhat strict |
|  |  | Family violence | Did your parents often quarrel? | often/sometimes |
|  |  |  | Have your father/mother ever beat up your mother/father? | often/sometimes |
|  |  | Parental separation or divorce | Were your biological parents divorced (including long separation due to emotional problems) before you were 17 years? | yes |
|  |  | Parental substance abuse | During the years you were growing up, which one of the followings did your male/female guardian ever have? | alcoholism/smoking/drug/gambling |
|  |  | Parents incarcerated | During the years you were growing up, which one of the followings did your male/female guardian ever have? | arrested/sent to prison |
|  |  | Parental mental illness | During the years you were growing up, had your male/female guardian showed continued signs of sadness or depression that lasted two weeks or more? | yes |
|  |  |  | Was this problem of your male/female guardian, sadness or depression during all, most, some, or only a little of your childhood? | all/most |
|  |  |  | Did your male/female guardian have abnormality of mind when you were young? | yes |
|  |  | Parental disability | Did your male/female guardian have a long time be sick on bed when you were young? | yes |
|  |  |  | Did your male/female guardian have a serious deformity when you were young? | yes |
|  |  | Parental death | Either of the parents was dead before participant was 17 years. | yes |
|  |  | Sibling death | Any of the siblings was dead before participant was 17 years. | yes |
|  |  | Physical abuse | When you were growing up, did your male/female guardian ever hit you? | often/sometimes |
|  |  |  | When you were growing up, how often did your brother or sister ever hit you? | often/sometimes |
|  |  | Economic adversity | When you were a child before age 17, compared to the average family in the same community/village at that time, how was your family’s financial situation? | a lot/somewhat worse off than them |
|  | Extra-familial | Bullying | When you were a child, how often were you picked on or bullied by kids in your neighborhood? | often/sometimes |
|  |  |  | When you were a child, how often were you picked on or bullied by kids in your school? | often/sometimes |
|  |  |  | When you were a child, how often did you feel worried about your physical safety at school? | often/sometimes |
|  |  | Loneliness | When you were a child, how often did you feel lonely for not having friends? | often/sometimes |
|  |  | Community violence | Was it safe being out alone at night in the neighborhood where you lived as a child? | not very safe/not safe at all |
|  |  |  | Were the neighbors of the place where you lived as a child very close-knit? | not very close-knit/not close-knit at all |
| HRS | Intra-familial | Emotional neglect | How much time and attention did your mother give you when you needed it? | a little/not at all |
|  |  |  | How much effort did your mother put into watching over you and making sure you had a good upbringing? | a little/not at all |
|  |  | Parental substance abuse | Before you were 18 years old, did either of your parents drink or use drugs so often that it caused problems in the family? | yes |
|  |  | Physical abuse | Before you were 18 years old, were you ever physically abused by either of your parents? | yes |
|  |  | Economic adversity | Would you say your family during that time was pretty well off financially, about average, or poor? | poor |
|  | Extra-familial | Repeating school year | Before you were 18 years old, did you have to do a year of school over again? | yes |
|  |  | Trouble with police | Before you were 18 years old, were you ever in trouble with the police? | yes |
| SHARE | Intra-familial | Emotional neglect | How much did your mother/father understand your problems and worries before you were age 17 | a little/not at all |
|  |  | Absent biological parent | Parents died before you were age 17 | yes |
|  |  |  | Didn't live with biological mother/father or had no female/male caregiver when ten | yes |
|  |  | Physical abuse | How often did your mother/ father push, grab, shove, throw something at you, slap or hit you before you were age 17? | often/sometimes |
|  |  | Economic adversity | Would you say your family during that time was pretty well off financially, about average, or poor from birth to age 16? | poor |
|  | Extra-familial | Non-parental abuse | How often did anybody else physically harm you in any way? | often/sometimes |
|  |  | Loneliness | Now I would like you to think back to your childhood, how often were you lonely for friends? | often/sometimes |

Abbreviations: ACEs, adverse childhood experiences; CHARLS, China Health and Retirement Longitudinal Study; HRS, Health and Retirement Study; SHARE, Survey of Health, Ageing and Retirement in Europe.

**eTable 2. Baseline Characteristics of included couples in the CHARLS, HRS, and SHARE**

| **Baseline Characteristics** | | **CHARLS** | |  | **HRS** | |  | **SHARE** | |
| --- | --- | --- | --- | --- | --- | --- | --- | --- | --- |
|  |  | **Husband (N=2394)** | **Wife (N=2394)** |  | **Husband (N=3131)** | **Wife (N=3131)** |  | **Husband (N=1831)** | **Wife (N=1831)** |
| Age, year | | 62.0 (57.0-68.0) | 60.0 (55.0-66.0) |  | 70.0 (61.0-77.0) | 66.0 (59.0-74.0) |  | 67.0 (61.0-73.0) | 64.0 (58.0-70.0) |
| Race | | | | | | | | | |
|  | White/Caucasian | - | - |  | 2617 (83.6) | 2643 (84.4) |  | - | - |
|  | Black or others | - | - |  | 514 (16.4) | 488 (15.6) |  | - | - |
| Residence | | | | | | | | | |
|  | Rural | 1512 (63.2) | 1512 (63.2) |  | - | - |  | 895 (48.9) | 895 (48.9) |
|  | Urban | 882 (36.8) | 882 (36.8) |  | - | - |  | 936 (51.1) | 936 (51.1) |
| Education | | | | | | | | | |
|  | Primary school or less | 1390 (58.1) | 1873 (78.2) |  | - | - |  | - | - |
|  | Middle school | 638 (26.7) | 362 (15.1) |  | - | - |  | - | - |
|  | High school or above | 366 (15.3) | 159 (6.6) |  | - | - |  | - | - |
|  | Lower than high school | - | - |  | 614 (19.6) | 514 (16.4) |  | - | - |
|  | High school | - | - |  | 850 (27.2) | 1010 (32.3) |  | - | - |
|  | College | - | - |  | 703 (22.5) | 843 (26.9) |  | - | - |
|  | above college | - | - |  | 964 (30.8) | 764 (24.4) |  | - | - |
|  | Education, years | - | - |  | - | - |  | 11.0 (10.0-13.0) | 11.0 (9.0-13.0) |
| Economic status | | | | | | | | | |
|  | Bottom tertile | 797 (33.3) | 797 (33.3) |  | 1042 (33.3) | 1042 (33.3) |  | 615 (33.6) | 615 (33.6) |
|  | Middle tertile | 797 (33.3) | 797 (33.3) |  | 1043 (33.3) | 1043 (33.3) |  | 595 (32.5) | 595 (32.5) |
|  | Top tertile | 800 (33.4) | 800 (33.4) |  | 1046 (33.4) | 1046 (33.4) |  | 621 (33.9) | 621 (33.9) |
| Smoking history | | | | | | | | | |
|  | Never smoking | 395 (16.5) | 2180 (91.1) |  | 1113 (35.6) | 1711 (54.7) |  | 928 (50.7) | 1432 (78.2) |
|  | Ever smoking | 1999 (83.5) | 214 (8.9) |  | 2018 (64.5) | 1420 (45.4) |  | 903 (49.3) | 399 (21.8) |
| Drinking history | | | | | | | | | |
|  | Never drinking | 875 (36.6) | 2119 (88.5) |  | 1164 (37.2) | 1381 (44.1) |  | - | - |
|  | Ever drinking | 1519 (63.5) | 275 (11.5) |  | 1967 (62.8) | 1750 (55.9) |  | - | - |
|  | No recent drinking | - | - |  | - | - |  | 1378 (75.3) | 1664 (90.9) |
|  | Recent drinking | - | - |  | - | - |  | 453 (24.7) | 167 (9.1) |
| BMI, kg/m^2^ | | 23.2 (20.8-25.7) | 24.3 (21.8-26.8) |  | 27.8 (25.1-31.2) | 27.4 (23.8-31.7) |  | 27.7 (25.0-30.6) | 27.6 (24.7-31.3) |
| WC, cm | | 86.3 (78.6-93.8) | 87.2 (80.0-94.5) |  | 104.8 (97.2-114.3) | 96.5 (86.4-106.7) |  | - | - |
| Hypertension | | | | | | | | | |
|  | No | 985 (41.1) | 1044 (43.6) |  | 933 (29.8) | 1108 (35.4) |  | 936 (51.1) | 936 (51.1) |
|  | Yes | 1409 (58.9) | 1350 (56.4) |  | 2198 (70.2) | 2023 (64.6) |  | 895 (48.9) | 895 (48.9) |
| Diabetes | | | | | | | | | |
|  | No | 2202 (92.0) | 2155 (90.0) |  | 2335 (74.6) | 2545 (81.3) |  | 1524 (83.2) | 1581 (86.4) |
|  | Yes | 192 (8.0) | 239 (10.0) |  | 796 (25.4) | 586 (18.7) |  | 307 (16.8) | 250 (13.7) |
| Dyslipidemia | | | | | | | | | |
|  | No | 2062 (86.1) | 2026 (84.6) |  | - | - |  | 1355 (74.0) | 1346 (73.5) |
|  | Yes | 332 (13.9) | 368 (15.4) |  | - | - |  | 476 (26.0) | 485 (26.5) |
| CVDs | | | | | | | | | |
|  | No | 2079 (86.8) | 1992 (83.2) |  | 2075 (66.3) | 2449 (78.2) |  | 1357 (74.1) | 1484 (81.1) |
|  | Yes | 315 (13.2) | 402 (16.8) |  | 1056 (33.7) | 682 (21.8) |  | 474 (25.9) | 347 (19.0) |
| Depressive symptoms* | | | | | | | | | |
|  | No | 1716 (75.8) | 1367 (62.1) |  | 2886 (93.1) | 2789 (89.4) |  | 1404 (78.6) | 1215 (67.3) |
|  | Yes | 547 (24.2) | 836 (38.0) |  | 213 (6.9) | 330 (10.6) |  | 382 (21.4) | 590 (32.7) |
| Overall ACEs | | | | | | | | | |
|  | 0 | 156 (6.5) | 225 (9.4) |  | 1239 (39.6) | 1495 (47.8) |  | 729 (39.8) | 747 (40.8) |
|  | 1 | 433 (18.1) | 510 (21.3) |  | 1050 (33.5) | 968 (30.9) |  | 569 (31.1) | 618 (33.8) |
|  | 2 | 547 (22.9) | 535 (22.4) |  | - | - |  | - | - |
|  | 2 or more | - | - |  | 842 (26.9) | 668 (21.3) |  | 533 (29.1) | 466 (25.5) |
|  | 3 | 484 (20.2) | 467 (19.5) |  | - | - |  | - | - |
|  | 4 or more | 774 (32.3) | 657 (27.4) |  | - | - |  | - | - |
| Intra-familial ACEs | | | | | | | | | |
|  | 0 | 187 (7.8) | 262 (10.9) |  | 1617 (51.6) | 1616 (51.6) |  | 846 (46.2) | 915 (50.0) |
|  | 1 | 496 (20.7) | 582 (24.3) |  | 1024 (32.7) | 990 (31.6) |  | 573 (31.3) | 594 (32.4) |
|  | 2 | 632 (26.4) | 595 (24.9) |  | - | - |  | - | - |
|  | 2 or more | - | - |  | 490 (15.7) | 525 (16.8) |  | 412 (22.5) | 322 (17.6) |
|  | 3 | 518 (21.6) | 466 (19.5) |  | - | - |  | - | - |
|  | 4 or more | 561 (23.4) | 489 (20.4) |  | - | - |  | - | - |
| Extra-familial ACEs | | | | | | | | | |
|  | 0 | 1688 (70.5) | 1743 (72.8) |  | 2223 (71.0) | 2761 (88.2) |  | 1471 (80.3) | 1408 (76.9) |
|  | 1 | 530 (22.1) | 493 (20.6) |  | 787 (25.1) | 353 (11.3) |  | 328 (17.9) | 384 (21.0) |
|  | 2 | - | - |  | 121 (3.9) | 17 (0.5) |  | 32 (1.8) | 39 (2.1) |
|  | 2 or more | 176 (7.4) | 158 (6.6) |  | - | - |  | - | - |

Abbreviations: CHARLS, China Health and Retirement Longitudinal Study; HRS, Health and Retirement Study; SHARE, Survey of Health, Ageing and Retirement in Europe; BMI, body mass index; WC, waist circumference; CVDs, cardiovascular diseases; ACEs, adverse childhood experiences. * means some of the recruited couples have missing data.

**eTable 3. Adjusted Spearman correlations of spousal multiple ACEs in the CHARLS, HRS, and SHARE**

|  | **Unadjusted** |  | **Model 1^a^** |  | **Model 2^b^** |  | **Model 3^c^** |
| --- | --- | --- | --- | --- | --- | --- | --- |
|  | **Spearman's correlation (95% CI)** | | | | | | |
| ***CHARLS*** | | | | | | | |
| Overall ACEs | 0.20 (0.16 to 0.23) |  | 0.19 (0.15 to 0.23) |  | 0.18 (0.14 to 0.22) |  | 0.18 (0.14 to 0.22) |
| Intra-familial ACEs | 0.17 (0.13 to 0.21) |  | 0.16 (0.12 to 0.20) |  | 0.16 (0.12 to 0.19) |  | 0.15 (0.11 to 0.19) |
| Extra-familial ACEs | 0.17 (0.13 to 0.21) |  | 0.17 (0.13 to 0.20) |  | 0.16 (0.13 to 0.20) |  | 0.16 (0.12 to 0.20) |
| ***HRS*** | | | | | | | |
| Overall ACEs | 0.14 (0.11 to 0.18) |  | 0.11 (0.07 to 0.14) |  | 0.10 (0.07 to 0.14) |  | 0.10 (0.07 to 0.14) |
| Intra-familial ACEs | 0.10 (0.06 to 0.13) |  | 0.08 (0.05 to 0.12) |  | 0.08 (0.04 to 0.11) |  | 0.08 (0.04 to 0.11) |
| Extra-familial ACEs | 0.10 (0.06 to 0.13) |  | 0.06 (0.03 to 0.10) |  | 0.06 (0.02 to 0.09) |  | 0.06 (0.02 to 0.09) |
| ***SHARE*** | | | | | | | |
| Overall ACEs | 0.29 (0.25 to 0.33) |  | 0.27 (0.23 to 0.31) |  | 0.27 (0.22 to 0.31) |  | 0.26 (0.22 to 0.31) |
| Intra-familial ACEs | 0.24 (0.19 to 0.28) |  | 0.21 (0.17 to 0.25) |  | 0.21 (0.16 to 0.25) |  | 0.21 (0.16 to 0.25) |
| Extra-familial ACEs | 0.27 (0.23 to 0.31) |  | 0.27 (0.23 to 0.31) |  | 0.26 (0.22 to 0.30) |  | 0.26 (0.22 to 0.30) |

Abbreviations: ACEs, adverse childhood experiences; CHARLS, China Health and Retirement Longitudinal Study; HRS, Health and Retirement Study; SHARE, Survey of Health, Ageing and Retirement in Europe; CI, confidence interval.

^a^ Model 1 was adjusted for both spouses’ age, race (HRS only), residence (CHARLS and SHARE only), education, and economic status.

^b^ Model 2 was further adjusted for both spouses’ body mass index, waist circumference (CHARLS and HRS only), smoking history, and drinking history based on Model 1.

^c^ Model 3 was further adjusted for both spouses’ diabetes, hypertension, dyslipidemia (CHARLS and SHARE only), and cardiovascular diseases based on Model 2.

**eTable 4. Association of respondents’ intra-familial and extra-familial ACEs with spousal depressive symptoms in the CHARLS, HRS, and SHARE^a^**

|  | **Husbands' ACEs on wives' depressive symptoms** | **Wives' ACEs on husbands' depressive symptoms** | |
| --- | --- | --- | --- |
|  | **OR (95% CI)** | | |
| ***The CHARLS*** | | | |
| No. of participants | 2203 | | 2263 |
| *Intra-familial* | | | |
| 0 | Reference | | Reference |
| 1 | 1.12 (0.75 to 1.68) | | 0.95 (0.65 to 1.40) |
| 2 | 1.37 (0.92 to 2.04) | | 1.09 (0.74 to 1.59) |
| 3 | 1.36 (0.91 to 2.04) | | 1.00 (0.67 to 1.48) |
| 4 or more | 1.90 (1.28 to 2.83) | | 1.56 (1.06 to 2.28) |
| Continuous | 1.15 (1.08 to 1.22) | | 1.09 (1.02 to 1.16) |
| *Extra-familial* | | | |
| 0 | Reference | | Reference |
| 1 | 0.94 (0.75 to 1.17) | | 1.05 (0.82 to 1.35) |
| 2 or more | 1.83 (1.30 to 2.59) | | 1.38 (0.94 to 2.02) |
| Continuous | 1.19 (1.05 to 1.36) | | 1.12 (0.96 to 1.30) |
| ***The HRS*** | | |  |
| No. of participants | 3119 | | 3099 |
| *Intra-familial* | | | |
| 0 | Reference | | Reference |
| 1 | 1.24 (1.07 to 1.44) | | 1.15 (0.98 to 1.34) |
| 2 or more | 1.23 (1.01 to 1.49) | | 1.04 (0.86 to 1.27) |
| Continuous | 1.12 (1.03 to 1.21) | | 1.04 (0.96 to 1.12) |
| *Extra-familial* | | | |
| 0 | Reference | | Reference |
| 1 | 1.05 (0.90 to 1.23) | | 1.06 (0.85 to 1.31) |
| 2 | 1.10 (0.78 to 1.55) | | 0.53 (0.20 to 1.44) |
| Continuous | 1.05 (0.93 to 1.19) | | 1.00 (0.82 to 1.23) |
| ***The SHARE*** | | |  |
| No. of participants | 1805 | | 1786 |
| *Intra-familial* | | | |
| 0 | Reference | | Reference |
| 1 | 1.11 (0.87 to 1.42) | | 1.06 (0.80 to 1.40) |
| 2 or more | 1.50 (1.15 to 1.96) | | 1.71 (1.24 to 2.36) |
| Continuous | 1.21 (1.08 to 1.35) | | 1.24 (1.09 to 1.40) |
| *Extra-familial* | | | |
| 0 | Reference | | Reference |
| 1 | 1.10 (0.83 to 1.45) | | 1.35 (1.01 to 1.81) |
| 2 | 1.45 (0.68 to 3.09) | | 3.08 (1.48 to 6.41) |
| Continuous | 1.13 (0.90 to 1.43) | | 1.48 (1.16 to 1.90) |

Abbreviations: ACEs, adverse childhood experiences; CHARLS, China Health and Retirement Longitudinal Study; HRS, Health and Retirement Study; SHARE, Survey of Health, Ageing and Retirement in Europe.

^a^ All ORs (95% CIs) between respondents’ (husbands' or wives') ACEs and spousal (wives' or husbands') depressive symptoms were adjusted for spousal ACEs, age, race (HRS only), residence (CHARLS and SHARE only), education, economic status, body mass index, waist circumference (CHARLS and HRS only), smoking history, drinking history, diabetes, hypertension, dyslipidemia (CHARLS and SHARE only), and cardiovascular diseases.

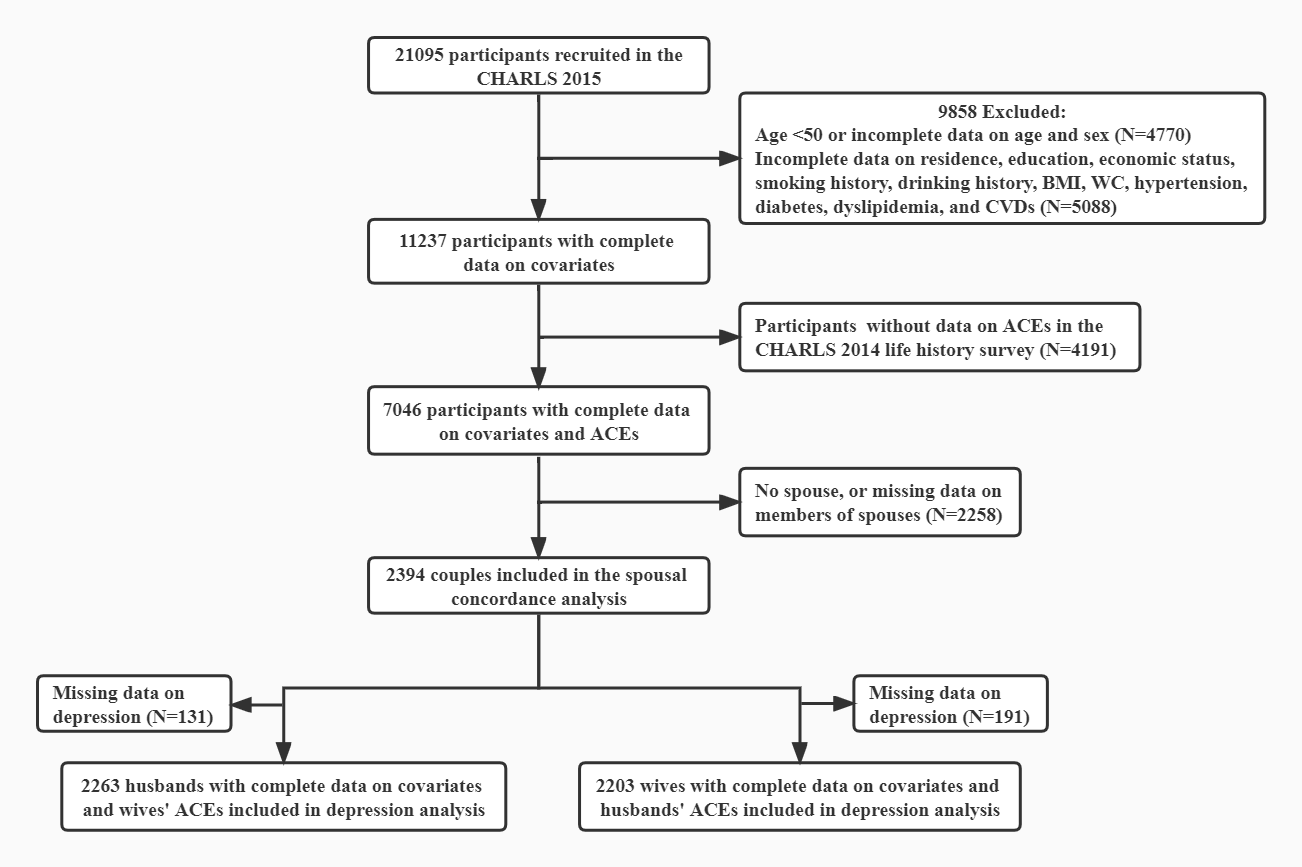

**eFigure 1. Flow chart of analyses in the CHARLS**

Abbreviations: CHARLS, China Health and Retirement Longitudinal Study; BMI, body mass index; WC, waist circumference; CVDs, cardiovascular diseases; ACEs, adverse childhood experiences.

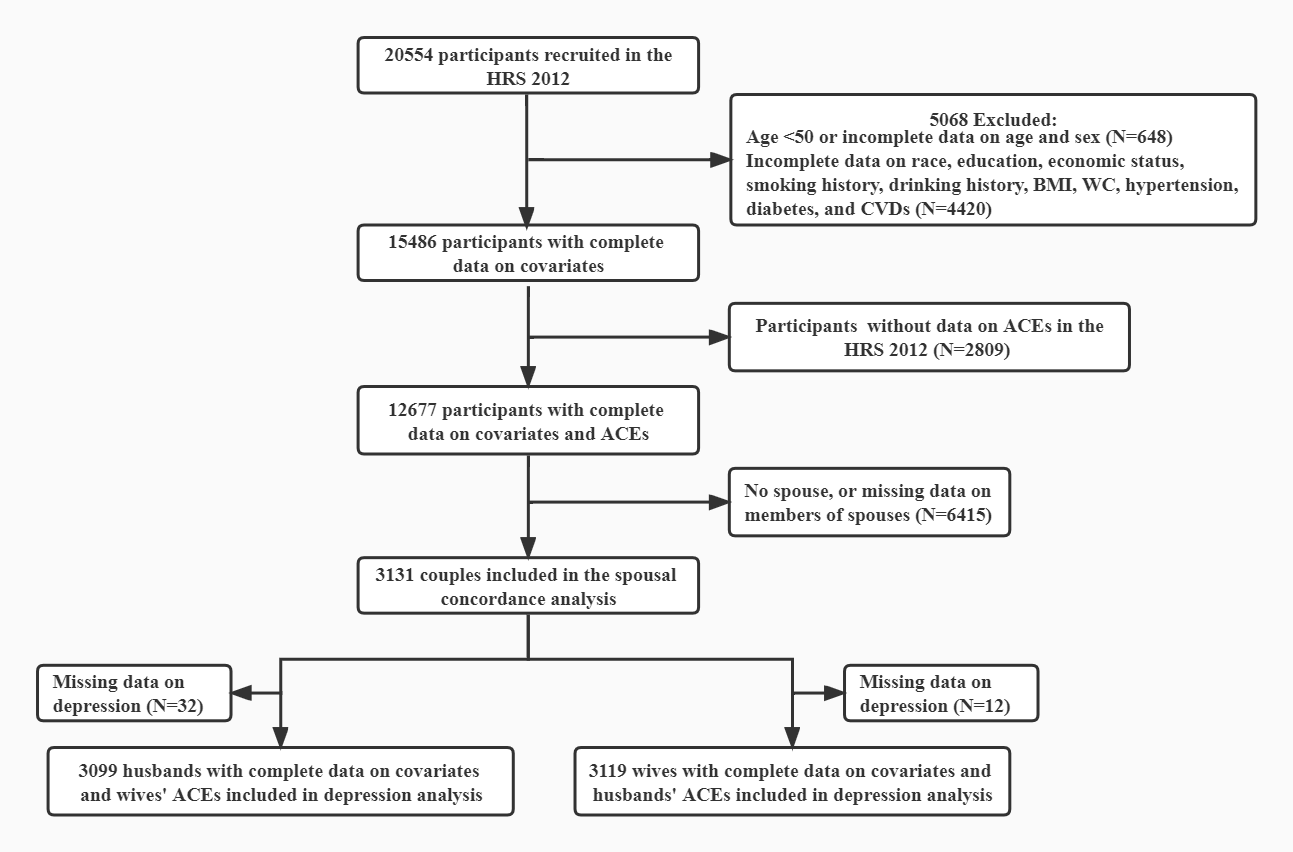

**eFigure 2. Flow chart of analyses in the HRS**

Abbreviations: HRS, Health and Retirement Study; BMI, body mass index; WC, waist circumference; CVDs, cardiovascular diseases; ACEs, adverse childhood experiences.

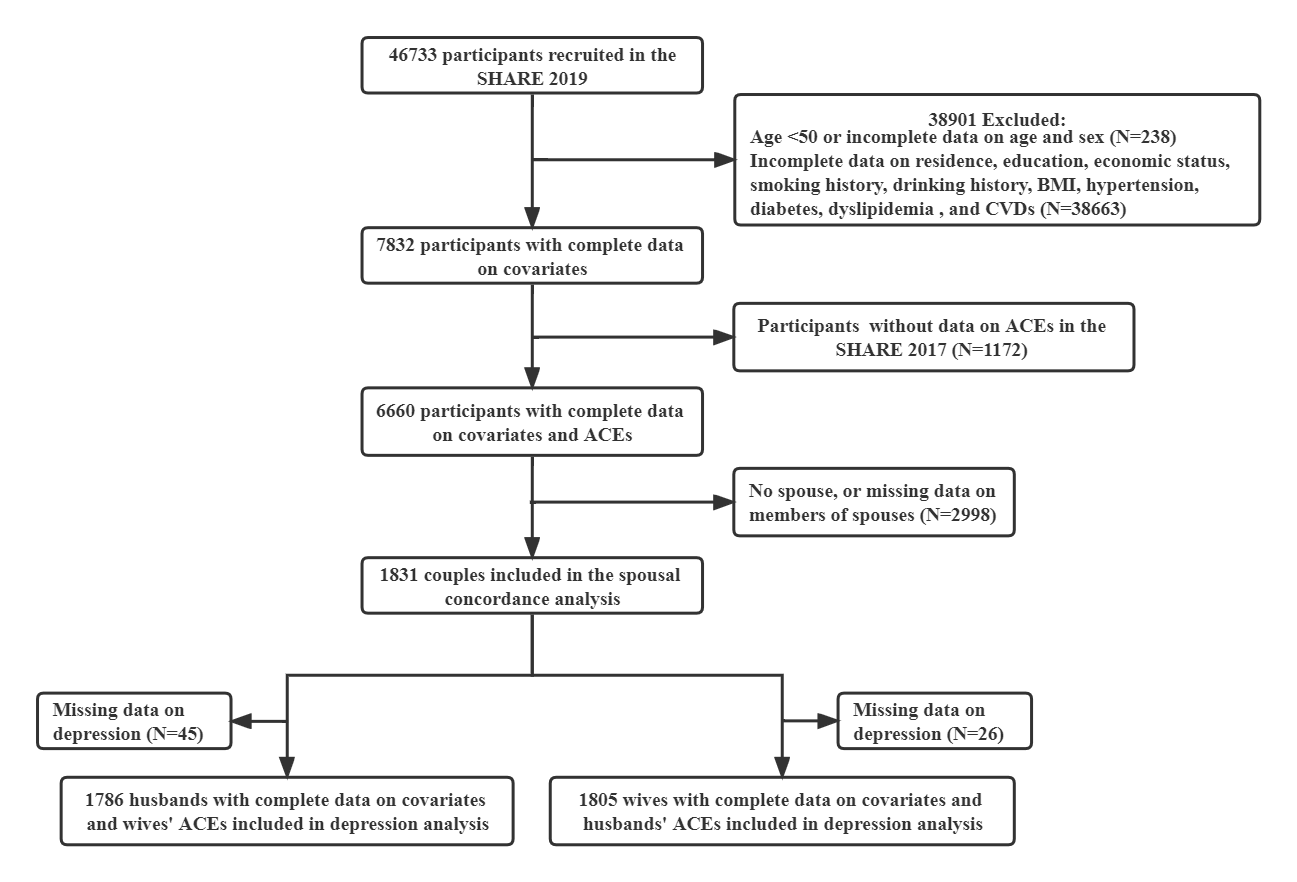

**eFigure 3. Flow chart of analyses in the SHARE**

Abbreviations: SHARE, Survey of Health; Ageing and Retirement in Europe; BMI, body mass index; CVDs, cardiovascular diseases; ACEs, adverse childhood experiences.
